# Determinants of Antibiotic Prescribing for Acute Respiratory Infections in Primary Care: A Retrospective Cross-Sectional Study at the Eduardo Mondlane University Health Center, Maputo, Mozambique

**DOI:** 10.64898/2026.09.05.26362349

**Authors:** Luís José Charanga, Avelino António Cuela

**Author notes:** **Correspondence:** Corresponding author: Luís José Charanga.

## Abstract

**Introduction:** Acute respiratory infections (ARIs) constitute one of the leading reasons for primary care consultations and, despite being frequently viral in etiology, continue to be inappropriately treated with antibiotics, contributing to the rise of antimicrobial resistance. In Mozambique, evidence on the clinical factors associated with antibiotic prescribing in patients with ARIs remains scarce, particularly in studies conducted in the general population.

**Objective:** To identify the clinical factors associated with antibiotic prescribing in patients with ARIs attended at the Eduardo Mondlane University Health Center (CS-UEM) between April and August 2024.

**Methods:** An observational, descriptive, cross-sectional, retrospective study with a quantitative approach, based on a convenience sample of 196 clinical records of patients with ARIs. Demographic, clinical, and prescriber-related variables were collected. Bivariate associations were assessed using chi-square (χ^2^) tests and Cramér’s V. A multivariable logistic regression model was subsequently fitted, whose discriminative capacity was evaluated using the area under the ROC curve (AUC).

**Results:** The prevalence of antibiotic prescribing was 69.4% (136/196), substantially higher than the range recommended by the World Health Organization (20–26.8%). Amoxicillin was the most frequently prescribed antibiotic (45.4%), followed by azithromycin (26.0%); 43.4% of prescriptions belonged to the WHO AWaRe “Watch” category. In the final multivariable model, clinical category and presenting signs and symptoms were independently and significantly associated with antibiotic prescribing (χ^2^(8) = 36.58; p < 0.001; McFadden’s pseudo-R^2^ = 0.151), whereas diagnosis, age, sex, and prescriber nationality showed no statistically significant association. The model demonstrated acceptable discriminative capacity (AUC = 0.760).

**Conclusion:** At CS-UEM, antibiotic prescribing for patients with ARIs appears to be determined chiefly by the clinical presentation at the time of consultation, rather than by the formal diagnosis or the patient’s demographic characteristics. These findings reinforce the need to implement antimicrobial stewardship interventions tailored to the local context, aimed at promoting the rational use of antibiotics.

## Introduction

Acute respiratory infections (ARIs) constitute a heterogeneous group of diseases affecting the upper and lower airways, characterized by sudden onset and a generally self-limited course, encompassing conditions such as the common cold, pharyngitis, sinusitis, otitis media, acute bronchitis, and pneumonia. Worldwide, these infections represent the leading cause of acute illness incidence and one of the most frequent reasons for seeking primary health care across all age groups [1]. Collectively, they contribute substantially to the global burden of morbidity, accounting for a considerable number of deaths and disability-adjusted life years, with a disproportionate impact on young children and other vulnerable populations [2]. Beyond the burden borne by individual patients, ARIs impose considerable strain on health systems, reflected in high rates of outpatient visits and diagnostic investigations, associated with direct and indirect economic costs arising from lost productivity and work and school absenteeism [1]. For these reasons, ARIs are widely recognized as a major public health problem requiring sustained attention from primary health care services.

A substantial proportion of ARIs, particularly those affecting the upper respiratory tract, are of viral origin, a circumstance in which antibiotic therapy confers no demonstrable clinical benefit [3]. Nevertheless, antibiotics continue to be frequently prescribed for these conditions in routine outpatient practice, reflecting a persistent gap between available scientific evidence and clinicians’ actual prescribing behavior [3]. Studies conducted in diverse settings show that the majority of ARI episodes still result in an antibiotic prescription, with a considerable proportion of these prescriptions issued in the absence of any recognized clinical indication [4]. Since the outpatient setting accounts for the largest share of global antibiotic consumption, understanding the determinants of prescribing at this level of care is of particular importance for antimicrobial stewardship efforts [4]. This observation is corroborated by a systematic review and meta-analysis of studies conducted in low- and middle-income countries, in which the prevalence of antibiotic prescribing in primary care ranged from 44% to 60%, with marked heterogeneity across settings [30]; a recent cross-sectional study conducted in eight primary care facilities in Zimbabwe reported an overall prescribing prevalence of 70.5%, close to that observed in the present study, with acute respiratory infections among the leading reasons for prescribing [31].

The inappropriate use of antibiotics carries consequences that extend far beyond the individual clinical episode. In 2019 alone, bacterial antimicrobial resistance is estimated to have been associated with nearly five million deaths worldwide, with the highest rates recorded in sub-Saharan Africa, the region where Mozambique is located [5,16,17]. A comprehensive review of antimicrobial resistance in sub-Saharan Africa identified limited access to health care, inappropriate antibiotic use, deficient surveillance, and weaknesses in pharmaceutical regulation as central determinants of this high regional burden [39]. Beyond its central role in the emergence and spread of resistance, unnecessary antibiotic use exposes patients to avoidable adverse drug reactions and adds further costs to health systems, related both to the acquisition of medicines and to the treatment of complications arising from their inappropriate use [4]. For these reasons, the World Health Organization (WHO) and various other international bodies have, in recent years, reinforced the need for more rational antibiotic use, placing this issue among today’s leading public health priorities.

The clinical decision to prescribe an antibiotic for an ARI is shaped by a broad range of characteristics related to the patient and the clinical presentation. Previous studies have identified associations between antibiotic prescribing and the presence of fever, productive cough, sore throat, and purulent rhinorrhea [6,11,12], as well as with prolonged symptom duration and abnormal physical examination findings, such as abnormal breath sounds or tonsillar hypertrophy [3]. Patient comorbidities and clinicians’ perception of disease severity have likewise been identified as relevant determinants of the prescribing decision [3]. However, the relative weight of each of these factors varies considerably across countries and levels of health care, suggesting that local, organizational, and cultural factors, beyond strictly clinical considerations, may significantly influence prescribing behavior in any given setting [3]. This contextual variability is illustrated by studies conducted in high-income health systems, in which the consultation modality (in-person vs. remote) influenced the likelihood of prescribing for ARIs [33], and by recent analyses of administrative data estimating that a substantial proportion of antibiotics prescribed for ARIs without a recognized indication occurs even in settings with broad access to health care [34].

Despite the global relevance of this issue, evidence on the clinical factors associated with antibiotic prescribing for ARIs in Mozambique remains limited, being largely confined to studies conducted in specific populations, such as HIV-positive patients followed in primary care [7]. More recent work, conducted in health facilities in the cities of Maputo and Matola, has sought to evaluate strategies to reduce unnecessary antibiotic prescribing in patients with upper respiratory tract infections, underscoring the continued relevance of this topic in the national context [8]. Studies on knowledge and practices regarding antibiotic use, among both health professionals and the community, conducted in Maputo and Manhiça, have likewise documented knowledge gaps and antibiotic use patterns that may favor inappropriate prescribing in Mozambique [22,23,24], within a national context recently described as an antimicrobial resistance crisis that cannot continue to be ignored [25]. Recent reviews on antibiotic prescribing in primary care in Africa emphasize that evidence on its clinical determinants remains fragmented and concentrated in a small number of countries, reinforcing the need for local studies in as-yet-undocumented settings, such as Mozambique [32]. To date, however, no published studies appear to have specifically evaluated the clinical factors associated with antibiotic prescribing for ARIs at the Eduardo Mondlane University Health Center, a facility serving a heterogeneous university and community population. This gap in the evidence limits the ability to design antimicrobial stewardship interventions tailored to the local context. Accordingly, this study aimed to identify the clinical factors associated with antibiotic prescribing in patients with acute respiratory infections attended at the Eduardo Mondlane University Health Center between April and August 2024.

## Methods

### Study design and setting

This study followed a descriptive, cross-sectional, retrospective design with a quantitative approach. It was conducted at the Eduardo Mondlane University Health Center (CS-UEM), a primary health care facility located in Maputo, Mozambique.

### Population and sampling

The study population comprised the clinical records of patients diagnosed with acute respiratory infections (ARIs) at CS-UEM between April and August 2024. A non-probabilistic convenience sample of 196 records was analyzed. The sample size was calculated assuming a 95% confidence level and an estimated antibiotic prescribing prevalence of 85%, based on values reported in prior literature. The antibiotic prescribing prevalence actually observed in the final sample (69.4%; 136/196) differed from this a priori estimate. This difference is expected, given that the assumed value was derived from studies conducted in different clinical and geographic settings, and does not compromise the validity of the sample, which remained adequate for detecting the associations examined in this study.

### Data collection

Data were extracted from paper-based clinical records using a structured data collection form. The variables collected included: demographic characteristics (age and sex); prescriber nationality (national or foreign); clinical presentation, recorded as the signs and symptoms presented; the clinical diagnosis assigned at the consultation; the clinical category to which each patient was allocated — an internal classification used at CS-UEM to group patients by service line according to the type of professional responsible for the initial visit: GPII (general practitioner), GN (general nurse), MCHN (Maternal and Child Health nurse), and S (stomatologist); the results of the auxiliary/complementary diagnostic examinations performed (chest X-ray, complete blood count, rapid SARS-CoV-2 test, and sputum smear microscopy); and the prescribing outcome, defined as whether or not an antibiotic was prescribed at the consultation.

### Statistical analysis

Data processing and statistical modeling were performed in Python 3.12. The pandas library was used for data cleaning and preparation, including merging the two original outcome indicators (“Prescription Yes” and “Prescription No”) into a single binary variable (1 = antibiotic prescribed; 0 = not prescribed), and correcting a data entry inconsistency identified in the sex variable. Descriptive statistics were then calculated, with frequencies and percentages for categorical variables and means with standard deviations (SD) for numerical variables, including age.

Bivariate associations between the clinical/demographic variables and the antibiotic prescribing outcome were assessed using chi-square (χ^2^) tests, performed with the scipy.stats library, with statistical significance set at p ≤ 0.05; the strength of significant associations was quantified using Cramér’s V. Age, sex, and prescriber nationality were included as candidate covariates in the full multivariable model, in order to control for potential demographic and prescriber-related confounders, consistent with the study’s aim of identifying independent clinical determinants of prescribing. Prior to multivariable modeling, associations between categorical predictors were examined pairwise using Cramér’s V, to identify potential collinearity. One level of the clinical category variable, corresponding to a small subgroup (n = 8) in which all patients had received an antibiotic, produced complete separation when included as a distinct category in the regression model; this level was therefore merged with a small, related category before model fitting.

A full multivariable logistic regression model was initially fitted, containing all candidate predictors, using the statsmodels library, with likelihood ratio tests used to assess the contribution of each variable. A parsimonious final model was then selected by comparing Akaike’s Information Criterion (AIC) and the Bayesian Information Criterion (BIC) between the full and reduced models, with adjusted odds ratios (ORs) and 95% confidence intervals obtained for this final model. The discriminative capacity of the final model was assessed using the sklearn.metrics module, used to generate a receiver operating characteristic (ROC) curve and calculate the area under the curve (AUC). Statistical significance was set at p ≤ 0.05 throughout all analyses.

## Results

### Data cleaning and analytic sample

A total of 196 patient records were available for analysis. Prior to statistical analysis, two data quality issues identified in the original database were corrected. First, the outcome had originally been recorded across two separate indicator columns (“Prescription Yes”/”Prescription No”), which were merged into a single binary variable (1 = antibiotic prescribed, 0 = not prescribed). Second, one record showed a data entry inconsistency in the sex variable (an erroneous leading space), which was corrected prior to analysis. After cleaning, no record had missing values for the variables of interest, so all 196 patients were retained in the final analytic sample.

### Descriptive characteristics

The mean age of participants was 27.8 years (SD = 22.2; median = 23.0; range 1–91), reflecting a wide age distribution spanning young children, adults, and older patients. Slightly more than half of participants were female (114/196; 58.2%). Overall, 136 of the 196 patients (69.4%) received an antibiotic prescription, while 60 (30.6%) did not. The distribution of clinical and demographic characteristics is presented in Table 1.

**Table 1.** Sociodemographic and clinical characteristics of study participants (n = 196)

| Variable | Category | n | % |
| --- | --- | --- | --- |
| Age, years — mean ± SD | 27.8 ± 22.2 (range 1–91) | 196 | — |
| Sex | Female | 114 | 58.2 |
|  | Male | 82 | 41.8 |
| Clinical category | GPll | 144 | 73.5 |
|  | GN | 31 | 15.8 |
|  | MCHN | 13 | 6.6 |
|  | S | 8 | 4.1 |
| Diagnosis | Pneumonia | 44 | 22.4 |
|  | Acute bronchitis | 42 | 21.4 |
|  | Bacterial laryngitis | 35 | 17.9 |
|  | Bacterial pharyngitis | 29 | 14.8 |
|  | Influenza | 25 | 12.8 |
|  | COVID-19 | 21 | 10.7 |
| Presenting signs/symptoms | Cough | 34 | 17.3 |
|  | Nasal congestion | 30 | 15.3 |
|  | Common-cold-type symptoms | 30 | 15.3 |
|  | Rhinorrhea | 28 | 14.3 |
|  | Fever | 27 | 13.8 |
|  | Other | 26 | 13.3 |
|  | Sore throat | 21 | 10.7 |
| Auxiliary examination | Chest X-ray | 57 | 29.1 |
|  | Complete blood count | 49 | 25.0 |
|  | Rapid SARS-CoV-2 test | 47 | 24.0 |
|  | Sputum smear microscopy | 43 | 21.9 |
| Prescriber nationality | National | 166 | 84.7 |
|  | Foreign | 30 | 15.3 |
| Antibiotic prescribed | Yes | 136 | 69.4 |
|  | No | 60 | 30.6 |

### Antibiotic prescribing patterns and comparison with WHO benchmarks

The overall antibiotic prescribing frequency observed (69.4%; 136/196) is approximately two to three times higher than the range recommended by the World Health Organization (20–26.8%) for outpatient prescribing quality [9]. Among the 136 patients who received an antibiotic, ten distinct antibiotics or antibiotic combinations were identified. Amoxicillin was the most frequently used agent, present in 45.4% of the total 196 patients (alone or in combination), followed by azithromycin (26.0%) and the amoxicillin/clavulanic acid combination (11.7%). Table 2 presents the frequency of each antibiotic and its corresponding WHO AWaRe classification, in its most recent version [10].

**Table 2.**
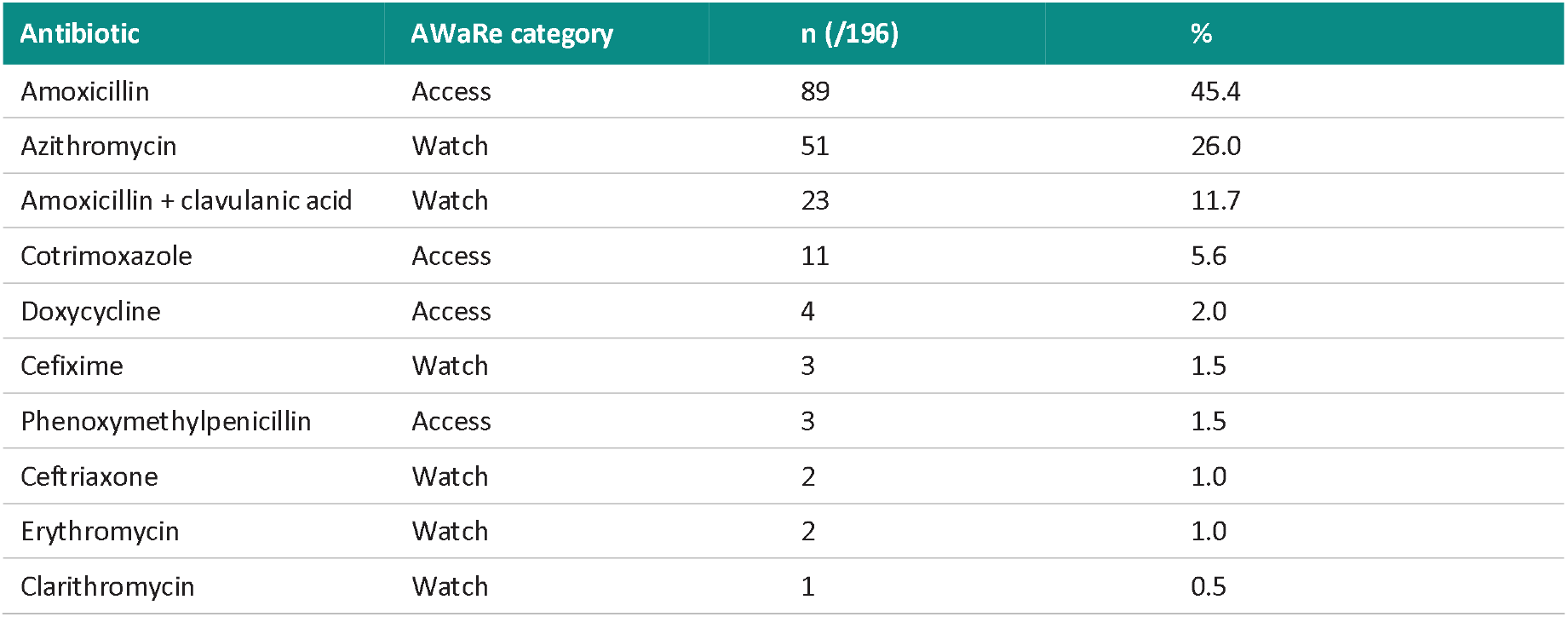
Frequency and WHO AWaRe classification of prescribed antibiotics (n = 196) Of the total 189 recorded antibiotic instances (i.e., counting each drug in a combination separately), 56.6% (107/189) belonged to the WHO “Access” category, considered first-line and lower risk in terms of resistance selection, while 43.4% (82/189) belonged to the “Watch” category, which requires closer monitoring due to greater potential for inducing antimicrobial resistance.

| Antibiotic | AWaRe category | n (/196) | % |
| --- | --- | --- | --- |
| Amoxicillin | Access | 89 | 45.4 |
| Azithromycin | Watch | 51 | 26.0 |
| Amoxicillin + clavulanic acid | Watch | 23 | 11.7 |
| Cotrimoxazole | Access | 11 | 5.6 |
| Doxycycline | Access | 4 | 2.0 |
| Cefixime | Watch | 3 | 1.5 |
| Phenoxymethylpenicillin | Access | 3 | 1.5 |
| Ceftriaxone | Watch | 2 | 1.0 |
| Erythromycin | Watch | 2 | 1.0 |
| Clarithromycin | Watch | 1 | 0.5 |

Regarding the number of antibiotics per prescription, 30.6% of patients (60/196) received no antibiotic, 42.3% (83/196) received a single antibiotic, and 27.0% (53/196) received a two-drug combination. Among the 53 combination prescriptions, the amoxicillin + azithromycin combination accounted for the large majority (96.2%; 51/53), with the remaining 3.8% (2/53) corresponding to the cotrimoxazole + ceftriaxone combination. Combination therapy was observed in both children (<16 years) and adults, with adults accounting for most combination prescriptions (30/53; 56.6%).

A Pareto analysis of the 189 total antibiotic instances (Figure 1) shows that the three most frequently used agents — amoxicillin, azithromycin, and amoxicillin/clavulanic acid — together accounted for 86.2% of all prescribed antibiotic instances, with amoxicillin and azithromycin alone already exceeding the conventional 80% threshold (74.1% cumulative). This concentration pattern, combined with the fact that two of these three leading agents belong to the WHO “Watch” category, indicates that antimicrobial stewardship interventions targeting a small number of specific therapeutic choices could cover most of the prescribing volume in this setting.

**Figure 1.**
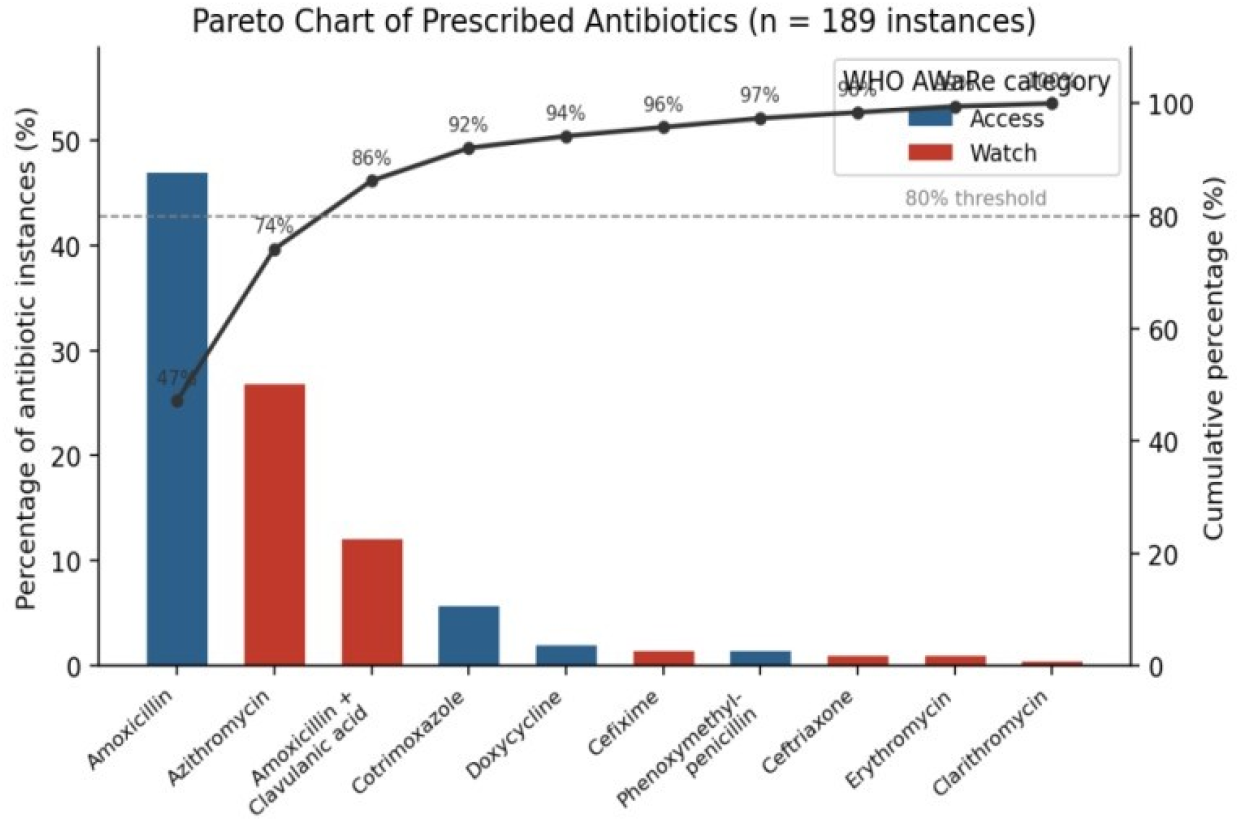
Pareto chart of prescribed antibiotics, showing individual frequency (bars, colored by WHO AWaRe category) and cumulative percentage (line) across the 189 recorded antibiotic instances (n = 196 patients).

It should be noted that the database used in this analysis recorded only the prescribed antibiotic(s), without a complete list of all medications issued per consultation. The core WHO prescribing indicators that require this broader information, such as the average number of medicines per prescription, the proportion of prescriptions with an injectable antibiotic, the proportion using generic naming, and compliance with the National Essential Medicines List, could therefore not be calculated from the available data and are not reported here.

### Bivariate associations with antibiotic prescribing

Chi-square tests of independence showed no statistically significant association between antibiotic prescribing and diagnosis, auxiliary examination, prescriber nationality, or patient sex. Patient age likewise did not differ significantly between those who received and those who did not receive an antibiotic (mean 27.1 vs. 29.4 years; Welch’s t = ™0.66; p = 0.512). In contrast, antibiotic prescribing was significantly associated with clinical category and presenting signs and symptoms, with Cramér’s V values indicating weak-to-moderate effect sizes for both associations (Table 3).

**Table 3.** Bivariate association between clinical/demographic variables and antibiotic prescribing.

| Variable | $\chi^2 / t$ | df | p-value | Cramér's V |
| --- | --- | --- | --- | --- |
| Sex | $\chi^2 = 0.19$ | 1 | 0.661 | 0.031 |
| Age (mean comparison) | $t = -0.66$ | — | 0.512 | — |
| Presenting signs/symptoms | $\chi^2 = 19.29$ | 6 | 0.00 | — |
| Diagnosis | $\chi^2 = 3.34$ | 5 | 0.648 | 0.131 |
| Auxiliary examination | $\chi^2 = 0.68$ | 3 | 0.877 | 0.059 |
| Clinical category | $\chi^2 = 15.45$ | 3 | 0.002 | 0.281 |
| Prescriber nationality | $\chi^2 = 0.32$ | 1 | 0.571 | 0.040 |

### Multivariable analysis

Prior to multivariable modeling, categorical predictors were examined for collinearity. Diagnosis showed moderate overlap with auxiliary examination (Cramér’s V = 0.436) and with clinical category (Cramér’s V = 0.396), and signs/symptoms overlapped moderately with diagnosis (V = 0.256) and with clinical category (V = 0.282), indicating that these variables capture partially overlapping clinical information. Additionally, one level of the clinical category variable (category “S”, n = 8) was associated with antibiotic prescribing in all eight cases; since this produced complete separation and an unstable, non-interpretable odds ratio when entered as a distinct level, this level was combined with the small “MCHN” category (combined n = 21) before fitting the model.

A multivariable logistic regression model including clinical category, signs/symptoms, diagnosis, age, and sex was statistically significant overall (likelihood ratio χ^2^(15) = 42.03; p < 0.001). Likelihood ratio tests for each block of variables showed that clinical category (χ^2^(2) = 14.25; p = 0.0008) and signs/symptoms (χ^2^(6) = 22.00; p = 0.0012) contributed significantly to the model, whereas diagnosis (χ^2^(5) = 5.37; p = 0.373), age (χ^2^(1) = 0.38; p = 0.538), and sex (χ^2^(1) = 0.05; p = 0.830) did not improve model fit after adjusting for the remaining variables.

A parsimonious final model was therefore fitted, including only clinical category and signs/symptoms. This reduced model achieved a lower (better) Akaike Information Criterion (AIC = 222.9 vs. 231.4) and Bayesian Information Criterion (BIC = 252.4 vs. 283.9) than the full model, confirming a more efficient fit without loss of explanatory power. The final model was statistically significant (likelihood ratio χ^2^(8) = 36.58; p < 0.001) and explained a modest but meaningful proportion of the variance in prescribing (McFadden’s pseudo-R^2^ = 0.151). The odds ratios of the final model are presented in Table 4.

**Table 4.**
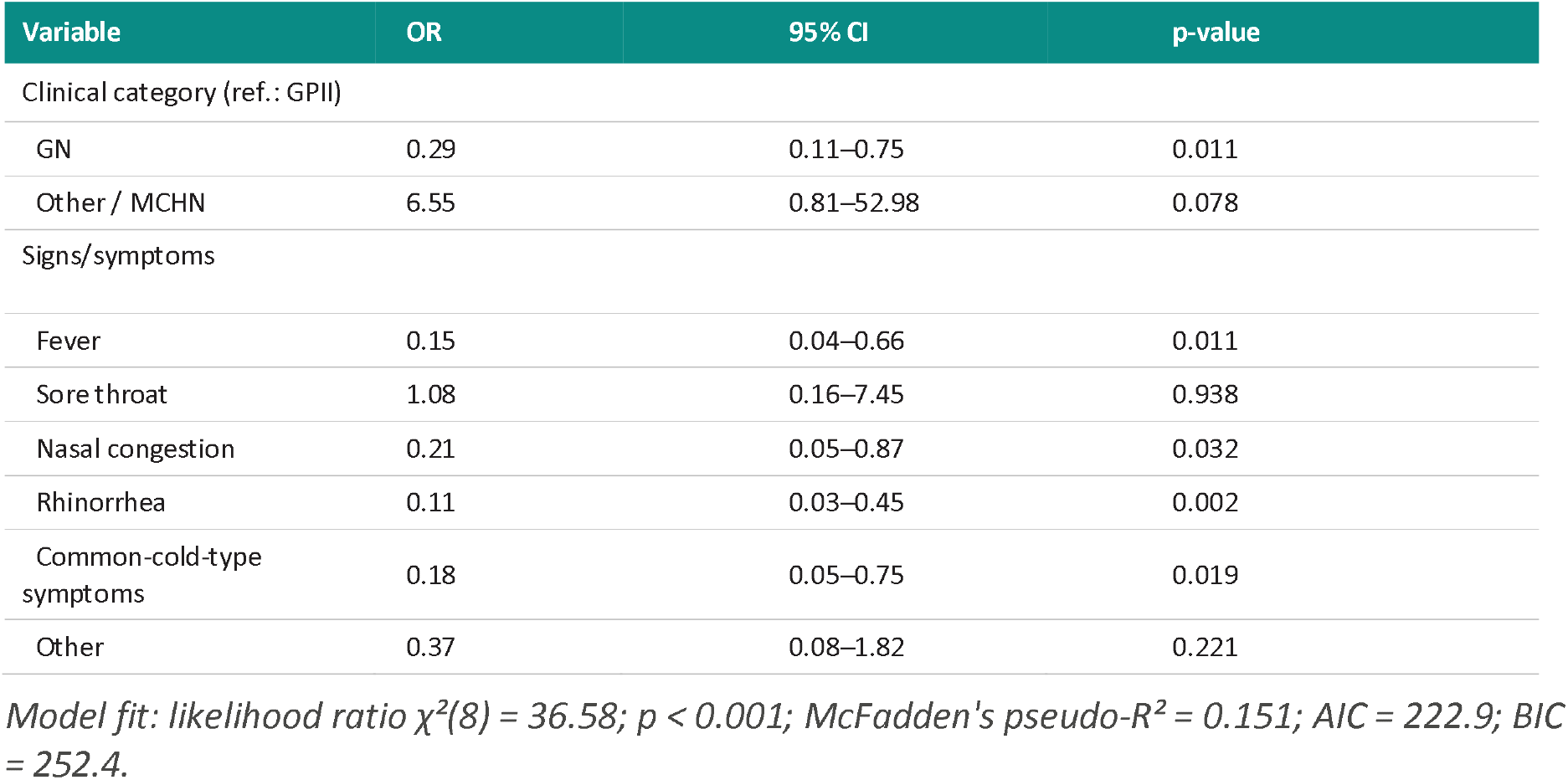
Final multivariable logistic regression model for antibiotic prescribing (n = 196) Compared with cough (reference category), fever, nasal congestion, rhinorrhea, and common-cold-type symptoms were each associated with significantly lower odds of antibiotic prescribing. Compared with the GPII clinical category, the GN category was associated with significantly lower odds of prescribing (OR = 0.29; 95% CI 0.11–0.75; p = 0.011) — that is, patients seen by a general nurse (GN) had, approximately, a 71% lower likelihood of receiving an antibiotic prescription than those seen by a general practitioner (GPII) — while the combined Other/MCHN category showed a positive association, corresponding to a likelihood of prescribing roughly 6.5 times higher than the GPII category, though statistically imprecise (OR = 6.55; 95% CI 0.81–52.98; p = 0.078), reflecting the small size of this subgroup.

### Model discrimination

The discriminative capacity of the final model was assessed through receiver operating characteristic (ROC) curve analysis. The model showed acceptable discrimination between patients who did and did not receive an antibiotic, with an area under the curve (AUC) of 0.760 (Figure 2), substantially higher than the value expected by chance (AUC = 0.500).

**Figure 2.**
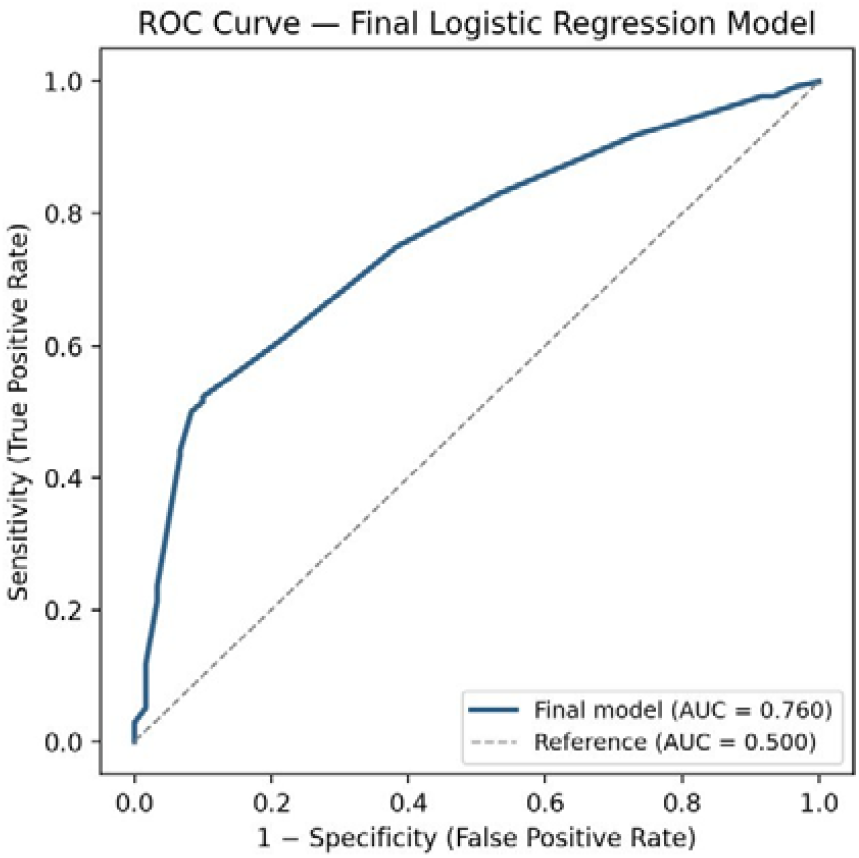
ROC curve for the final multivariable logistic regression model for antibiotic prescribing (predictor variables: clinical category and presenting signs/symptoms; AUC = 0.760), illustrating acceptable discriminative capacity between patients with and without a prescription.

### Summary of results

In summary, after correcting the data entry inconsistencies in the outcome and sex variables, and coding the multicategorical clinical variables as indicator (dummy) contrasts rather than ordinal numeric codes, clinical category and presenting signs and symptoms emerged as independent, statistically significant correlates of antibiotic prescribing, whereas diagnosis, patient age, and sex were not. The final model showed a statistically significant improvement over the null model (p < 0.001) and acceptable discriminative capacity (AUC = 0.760), indicating that clinical presentation variables meaningfully, though not fully, explain prescribing patterns in this population

## Discussion

This study examined the clinical factors associated with antibiotic prescribing in patients with acute respiratory infections attended at the Eduardo Mondlane University Health Center. After data validation, correction of outcome coding, appropriate coding of multicategorical clinical variables, and explicit handling of a separation issue in one level of the clinical category variable, the analysis showed that presenting signs and symptoms and the assigned clinical category were independently and significantly associated with the decision to prescribe an antibiotic, whereas the specific diagnosis, patient age, and sex were not. The final model reasonably discriminated between patients who did and did not receive an antibiotic (AUC = 0.760), indicating that clinical presentation, rather than diagnosis or demographic characteristics, was the main correlate of prescribing behavior in this setting. It should be noted, however, that the final model’s McFadden’s pseudo-R^2^ (0.151) was modest, indicating that a substantial portion of the variation in antibiotic prescribing remains unexplained, likely related to factors not measured in this study, such as symptom duration, perceived disease severity, or prescriber characteristics.

The fact that signs and symptoms outweighed diagnosis as a determinant of prescribing is consistent with a substantial body of international literature. Clinical judgment analysis studies have shown that prescribing decisions for respiratory infections are strongly influenced by the presence of specific signs, such as fever, sore throat, and purulent secretions, largely independent of the underlying diagnosis [6,15], even in primary care settings outside Mozambique [11,12]. Similarly, a systematic review of factors associated with antibiotic prescribing for respiratory tract infections concluded that physical examination findings were consistently stronger predictors of prescribing than the diagnostic label itself, and that the relative influence of these findings varied considerably across settings [3,12,13]. Similar observations have been described in high-income settings, where the clinical presentation and the consultation format itself, not just the diagnosis, were associated with the prescribing decision [33,34]. Our results are consistent with this pattern and suggest that similar prescribing behavior may also occur in this Mozambican primary care setting, in which prescribing appears to be guided mainly by the clinical picture at presentation, rather than by a formal diagnostic classification.

The absence of an independent association between diagnosis and antibiotic prescribing is a notable finding, considering that the diagnoses recorded in this cohort differ substantially in their expected bacterial etiology, ranging from conditions such as influenza and COVID-19, for which antibiotics are not indicated, to conditions such as pneumonia and bacterial pharyngitis, for which they may be warranted. The absence of an independent association indicates that, after adjusting for the other variables included in the model, the recorded diagnosis did not add further explanatory information regarding antibiotic prescribing. This finding is consistent with reports from other settings, in which a considerable proportion of antibiotics prescribed for acute respiratory infections are issued even when the recorded diagnosis does not support their use [4], a pattern also identified in a meta-analysis of primary care studies in low and middle-income countries [30]. Combined with the moderate collinearity observed between diagnosis, clinical category, and auxiliary examination in this dataset, these findings suggest that clinicians may rely more on the clinical features of presentation than on the recorded diagnostic label when making prescribing decisions, although the overlap among these clinical variables may also have reduced the independent contribution of diagnosis in the multivariable model.

The absence of an association between patient age, sex, or prescriber nationality and antibiotic prescribing suggests that, in this setting, prescribing decisions were not strongly influenced by the patient’s demographic characteristics, nor by whether the prescriber was of national or foreign origin. This is a reassuring observation from an equity standpoint, although it should be interpreted with caution, given the relatively small proportion of foreign prescribers and older patients in the sample, which may have limited the statistical power to detect smaller-magnitude effects.

From a public health perspective, these findings have direct relevance for antimicrobial stewardship. Given that a substantial proportion of the respiratory infections observed in primary care are viral and self-limited in origin, and that unnecessary antibiotic prescribing remains one of the main drivers of the antimicrobial resistance burden disproportionately affecting sub-Saharan Africa [5,16,17,18], the observation that antibiotic prescribing was more strongly associated with presenting signs and symptoms than with diagnosis suggests that structured interventions such as the clinical decision support algorithm already tested in HIV-positive patients with upper respiratory tract infections in Maputo and Matola [8], together with other antimicrobial stewardship initiatives already implemented or proposed in other countries in the region [19,20,21], including community-based interventions targeting both providers and communities in West and Central Africa [38], pharmacist-led antimicrobial stewardship programs [37], and analyses of the implementation determinants of these programs in resource-limited settings [36] could usefully be extended to the broader population of outpatients attended at the Eduardo Mondlane University Health Center. Incorporating diagnosis-specific prescribing criteria into routine practice, together with targeted training on the limited value of nonspecific signs and symptoms as indicators of bacterial infection, could help better align prescribing with the actual clinical indication.

An additional point worth highlighting is the antibiotic use pattern observed in this study, with nearly half of prescriptions involving WHO AWaRe “Watch” category antibiotics namely azithromycin and the amoxicillin/clavulanic acid combination and a notable proportion of combination prescriptions, especially the amoxicillin + azithromycin combination. This pattern is in line with concerns identified in other African settings, where the proportion of Watch antibiotics prescribed in outpatient care frequently remains above the WHO target of at least 60–70% Access category consumption [10,28,29]. A comparable pattern was recently described in a large cross-sectional study in primary care in Zimbabwe, in which amoxicillin was likewise the most frequently prescribed antibiotic, but about a third of prescriptions belonged to the Watch category [31], and recent reviews of antibiotic prescribing in Africa identify this deviation from WHO targets as a challenge common across the region, requiring antimicrobial stewardship interventions tailored to each context [32]. The preferential use of azithromycin, in particular, is a further cause for concern given its broad spectrum of activity and its well-documented role in selecting for macrolide resistance, such that monitoring and reducing its empirical use for ARIs without confirmed bacterial indication should be a priority target of any antimicrobial stewardship intervention implemented at CS-UEM.

This study has several strengths, including the use of routinely collected clinical records, the correction of data quality issues that would otherwise have obscured genuine associations, and the use of an analytic approach appropriate to categorical clinical data, including explicit handling of near-complete separation. Nonetheless, several limitations must be acknowledged. First, the retrospective, single-center design and the relatively modest sample size (n = 196) limited statistical power, particularly for less frequent categories, and restricted the number of predictors that could be reliably estimated, this may partly explain why diagnosis and demographic variables did not reach statistical significance despite their clinical plausibility. The single-center nature of the study also limits the external validity of the results: since CS-UEM is a university-affiliated primary care facility, with a service-line organization and a prescriber composition that may not be representative of other Mozambican health facilities, the prescribing patterns described here should be generalized cautiously to settings with different resource levels, clinical staff composition, or local protocols, and multicenter studies are needed to assess their broader applicability. Related to this point, the clinical interpretation of the clinical category variable (GPII, GN, MCHN, S) remains limited until the exact criteria used to allocate patients to each of these categories at CS-UEM are made explicit, without this clarification from the service, the finding that the GN category is associated with a lower likelihood of prescribing (Table 4) cannot be fully interpreted or replicated by other researchers. Second, the database did not include symptom duration, disease severity, or laboratory markers such as C-reactive protein, all previously identified as relevant predictors of prescribing [3,6], so residual confounding by unmeasured clinical factors cannot be excluded. Third, information at the prescriber level, such as years of clinical experience, and at the patient level, such as expressed expectations regarding antibiotic treatment, was not available, both of which have previously been shown to influence prescribing decisions [3]. Additionally, since the study was based on routinely collected clinical records, some degree of incomplete or inconsistent clinical documentation cannot be excluded, a limitation also noted in other reviews based on routine data from African health systems [35]. Similar patterns of antibiotic prescribing driven by clinical presentation, at times in excess of what protocols indicate, have also been described in other pediatric and outpatient care settings in low and middle-income countries [26,27], reinforcing the external plausibility of this study’s findings. Finally, as this is an observational study, these findings describe associations and do not establish a causal relationship between clinical presentation and prescribing.

## Conclusion

Antibiotic prescribing for acute respiratory infections at the Eduardo Mondlane University Health Center, between April and August 2024, was independently associated with presenting signs and symptoms and with clinical category, but not with diagnosis, age, sex, or prescriber nationality. These findings suggest that prescribing in this context appears to be predominantly guided by the clinical presentation at the time of care, rather than by a formal diagnostic or demographic profile, reinforcing the need for locally adapted antimicrobial stewardship interventions targeting the specific signs and symptoms most strongly associated with unjustified prescribing

## Data Availability

All data produced in the present study are available upon reasonable request to the authors

## Abbreviation Meaning

Table 5 List of acronyms and abbreviations used in the manuscript

ARI: Acute Respiratory Infection(s)
WHO: World Health Organization
CS-UEM: Eduardo Mondlane University Health Center
HIV: Human Immunodeficiency Virus
AWaRe: Access, Watch, Reserve (WHO) antibiotic classification
SD: Standard Deviation
95% CI: 95% Confidence Interval
OR: Odds Ratio
χ^2^: Chi-square test statistic
df: Degrees of freedom
AIC: Akaike Information Criterion
BIC: Bayesian Information Criterion
ROC: Receiver Operating Characteristic
AUC: Area Under the Curve
MCHN: Maternal and Child Health nurse
GPII: General practitioner
GN: General nurse
S: Stomatologist

## Acknowledgements

The authors thank the Eduardo Mondlane University Health Center for the authorization granted to conduct this study.

## Author contributions

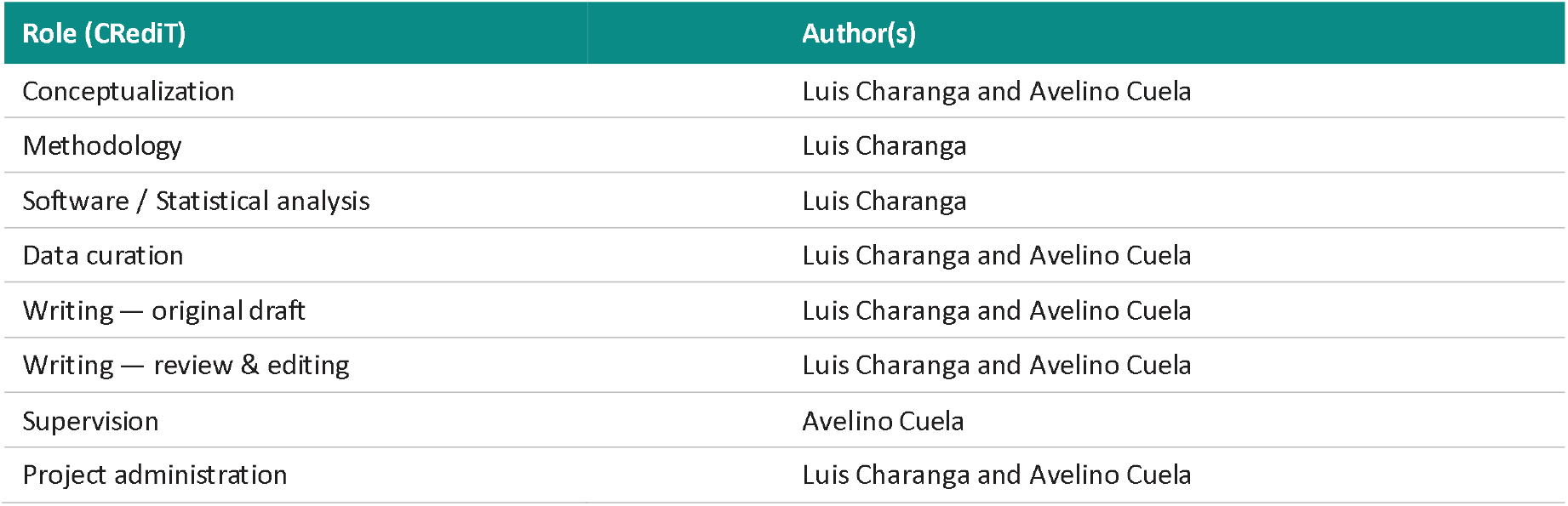

All authors reviewed and approved the final version of the manuscript submitted for publication.

## Funding

This study did not receive specific funding from public, commercial, or not-for-profit funding agencies.

## Data availability

The data supporting the findings of this study were extracted from clinical records of the Eduardo Mondlane University Health Center and are not publicly available, owing to patient confidentiality considerations. Anonymized data may be made available by the corresponding author upon reasonable and justified request, subject to approval by the institution responsible for the data. The structured data collection form and the Python code used for data cleaning and statistical modeling can be made available as supplementary material upon request.

## Declarations

### Ethics approval and consent to participate

This study was based on the retrospective review of clinical records and therefore did not involve direct contact with patients or any intervention beyond routine clinical care. The study protocol was conducted in accordance with the ethical principles of the Declaration of Helsinki and was approved by the Institutional Health Bioethics Committee of the Faculty of Medicine/Maputo Central Hospital (CIBS/FM&HCM), under reference number CIBS/FM&HCM/104/2024. As this involved retrospective, anonymized data collection with no direct patient interaction, the CIBS/FM&HCM waived the requirement for individual informed consent. All data were anonymized prior to statistical analysis, precluding individual patient identification.

### Consent for publication

Not applicable — the manuscript does not contain individual data or images that would permit patient identification.

### Competing interests

The authors declare that they have no competing financial or personal interests that could inappropriately influence the work presented in this article.

### Declaration of Generative AI and AI-Assisted Technologies in the Writing Process

During the preparation of this manuscript, the authors used Claude.ai to assist with the formatting and organization of the text and Grammarly to assist with grammar checking. The authors reviewed and verified the suggestions provided by these tools and take full responsibility for the content of the publication.

## Notes

### Competing Interest Statement

The authors have declared no competing interest.

### Summary of Updates

There are no significant changes to the scientific content of the manuscript. This version includes corrections to pagination and table formatting to improve the overall presentation, readability, and consistency of the manuscript.

